# Neuropathology-based GWAS for Alzheimer’s disease reveals novel susceptibility loci and highlights sex-specific pathways

**DOI:** 10.1101/2023.12.17.23300106

**Authors:** Yin Jin, Apostolia Topaloudi, Sudhanshu Shekhar, Alicia Nicole Scott, Bryce David Colon, Petros Drineas, Chris Rochet, Peristera Paschou

## Abstract

Dementia refers to an umbrella phenotype of many different underlying pathologies with Alzheimer’s disease (AD) being the most common type. While Genome-wide Association Studies (GWAS) of clinically defined AD have identified multiple AD susceptibility variants, a significant portion of the heritability remains unexplained highlighting the phenotypic and genetic heterogeneity of the clinically defined entity. Furthermore, despite women’s increased susceptibility to dementia there is a lack of sex-specific genetic studies and understanding of sex-specific background for the disorder. Here, we aim to tackle the heterogeneity of AD by specifically concentrating on neuropathological features which are the gold standard for diagnosis and pursuing sex-specific analysis. We bring together 13 different genomic and neuropathology datasets (6,960 individuals) and we integrate our GWAS findings with transcriptomic and proteomic data (ROSMAP and Mayo studies) aiming to also identify in vivo biomarkers for AD progression. We uncover novel genetic associations to AD neuropathology, including BIN1, OPCML, and CDH4. Our sex-specific analysis points to a role for BIN1 specifically in women as well as novel AD loci including QRFPR and SGCZ. Tissue-specific associations in females, particularly in the ovary issue, suggest a connection between sex hormones and AD. Finally, we pursue the identification of in-vivo biomarkers associated with AD neuropathology, with a goal to offer insights into disease progression and potential therapeutic targets. Our findings contribute to unraveling the molecular basis of AD, emphasizing the importance of sex-specific analyses and multi-omics approaches. Further research is needed to validate and explore the clinical utility of these findings.

## Introduction

Dementia, characterized as a persistent acquired disorder of mental processes involving memory problems, personality shifts, and impaired reasoning, ranks among the most prevalent age-related illnesses worldwide and is associated with great public health burden and costs ^1^. While Alzheimer’s disease (AD) is the most widespread form, other types of dementia, like vascular dementia, Lewy body dementia, and frontotemporal dementia also exist, sharing common clinical features. Thus, accurate clinical diagnosis of the specific type of dementia is challenging because multiple pathologies can give rise to similar clinical syndromes ^2,3^. In fact, accurate AD diagnosis can only be placed based on neuropathological findings and AD assessed through cognitive function, carries an approximate 24% misdiagnosis rate, especially when dealing with mixed dementia phenotypes ^4^. This heterogeneity also hampers genetic studies with a large portion of AD heritability still unexplained ^5^. Furthermore, even though women carry a higher risk of developing dementia, genomic studies that investigate sex-specific genetic background in dementia are lacking. Undoubtedly, this phenotypic and sex-related heterogeneity must be tackled to elucidate the genetic basis and neurobiology of dementia and drive accurate diagnosis and effective patient management.

Genome-wide Association Studies (GWAS) have led to the identification of more than 20 AD susceptibility common variants and rare genetic factors ^6^. However, AD GWAS based on clinical AD diagnosis are hampered by inclusion of pre-clinical patients in the control group and the use of a pathologically heterogeneous disease phenotype^7^. Furthermore, the extent to which identified variants are risk factors for AD pathology, coexisting pathologies, or other neurobiologic indices is unclear ^8,9^ limiting the potential use of GWAS findings to guide drug design for AD and inform clinical trials. In fact, Farfel et al ^10^, showed that many recently discovered genomic variants for AD dementia are not associated with the pathology of AD. Through neuropathological examinations, such as post-mortem brain analyses, it is possible to uncover distinctive brain abnormality patterns associated with AD, marked by the presence of neuritic plaques (NPs) and tau neurofibrillary tangles (NFTs). Demonstrating the power of this approach and despite using a much smaller sample than traditional GWAS, Beecham et al ^11^ were able to confirm association to APOE to common AD pathologies. There is thus a need to further extend GWAS based on neuropathologically-confirmed AD.

Another factor to take into consideration in AD genomic studies is the importance of studying sex-specific differences given the observed prevalence and progression of the disease in men versus women. Women are more likely to develop AD than men, and they also exhibit more tau protein tangles in their brains, leading to faster cognitive decline compared to men ^12^. Dumitrescu et al ^13^ performed a sex-stratified GWAS on AD neuropathology measurements in a sample of 2,701 males and 3,275 females, the majority of whom were diagnosed with AD at autopsy. They found that, outside of the APOE region, one locus on chromosome 7 (rs34331204) showed a sex-specific association with neurofibrillary tangles among males but not females, implicating a novel locus that confers male-specific protection from tau pathology. Such studies highlight the value of assessing genetic associations in a sex-specific manner.

Although post-mortem studies ensure an accurate AD diagnosis and can help tackle the heterogeneity of AD, in-vivo biomarkers are needed in order to move towards risk prediction or early diagnosis and early intervention that would prevent or delay symptoms. Recent work has shown that dementia-associated pathological changes may start 20-30 years before clinical onset ^14–16^ and newer AD drugs are being tested on pre-symptomatic participants aiming to halt or slow down cognitive decline before substantial damage has been done to the brain. There is thus an urgent need to predict early pre-symptomatic individuals as well as aim to differentiate towards specific neuropathology that leads to cognitive decline.

Here, we present a large-scale integrative analysis investigating the genomic background of neuropathologically-confirmed AD (ncAD) as well as continuous measures of Braak stage and NP score. Through neuropathology-based GWAS, sex-specific analysis and multiomic approaches we aimed to help disentangle the heterogeneous nature of the dementia clinical phenotype and move towards targeted therapies. Our work uncovered novel genes that underlie AD neuropathology, target genes with potential causal effects on disease progression, and presents a framework towards the identification of in-vivo biomarkers for AD.

## Results

### Genome-wide association studies of AD neuropathology

First, we conducted GWAS meta-analyses for three phenotypes: neuropathologically-confirmed AD (ncAD) on a total of 5,384 cases and 1,576 controls, Braak stage, and NP stage on 6,960 individuals, integrating 13 large-scale genetic, clinical, and neuropathology datasets from multiple sources (see Methods as well as Supplementary Materials, Supplementary Table S1). We identified two genomewide significant loci associated to AD neuropathology (Table 1). The top and only locus shared by all three GWAS was 19q13.32 near the APOE region (spanning TOMM40, APOE, PVRL2, BCAM, and APOC1). SNP rs184017 (p-value=5.8×10^−55^, Z=-9.55) was the top SNP in the case-control ncAD GWAS (Table 1, Figure 1A), while rs34342646 was the top SNP in both Braak stage (p-value=9.85×10^−53^, Z=15.28) (Table 1, Figure 2C) and rs34095326 was the top SNP in NP score (p-value=3.91×10^−34^, Z=12.18) GWAS (Table 1, Figure 1E). The second genome-wide significant locus shared by case-control ncAD GWAS (top SNP: rs4663105, p-value=7.94×10^−9^, Z=-5.77) (Table 1, Figure 1A, Supplementary Figure 1A) and Braak stage (top SNP:rs6733839, p-value=3×10^−8^, Z=5.54) was on chromosome 2q14 on the BIN1 gene (Table 1, Figure 1C, Supplementary Figure 2B).

**Figure 1:**
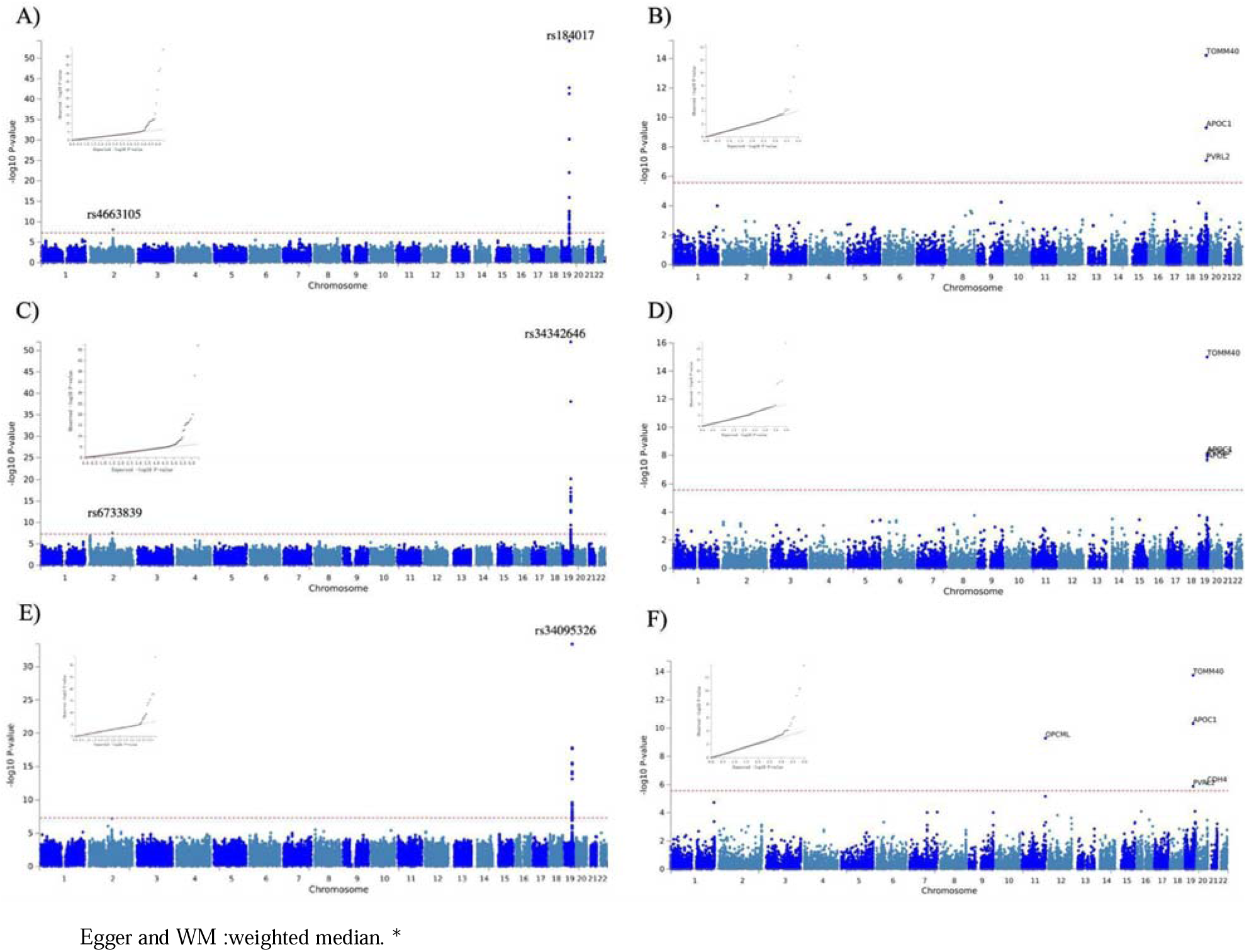
Manhattan and QQ plots of SNP-based and Gene-Based genome-wide association results of neuropathological features of AD (n□=□6,960). Dotted red lines represent the threshold for genome-wide significance (P <5×10^−8^) and Bonferroni correction for the gene-based analyses. A) Neuropathologically-confirmed AD case-control GWAS. B) Gene-based analysis for neuropathologically-confirmed AD case-control sample. C) Braak stage GWAS. D) Gene-based analysis for Braak stage E) NP score GWAS. F) Gene-based analysis for NP score.

**Figure 2.**
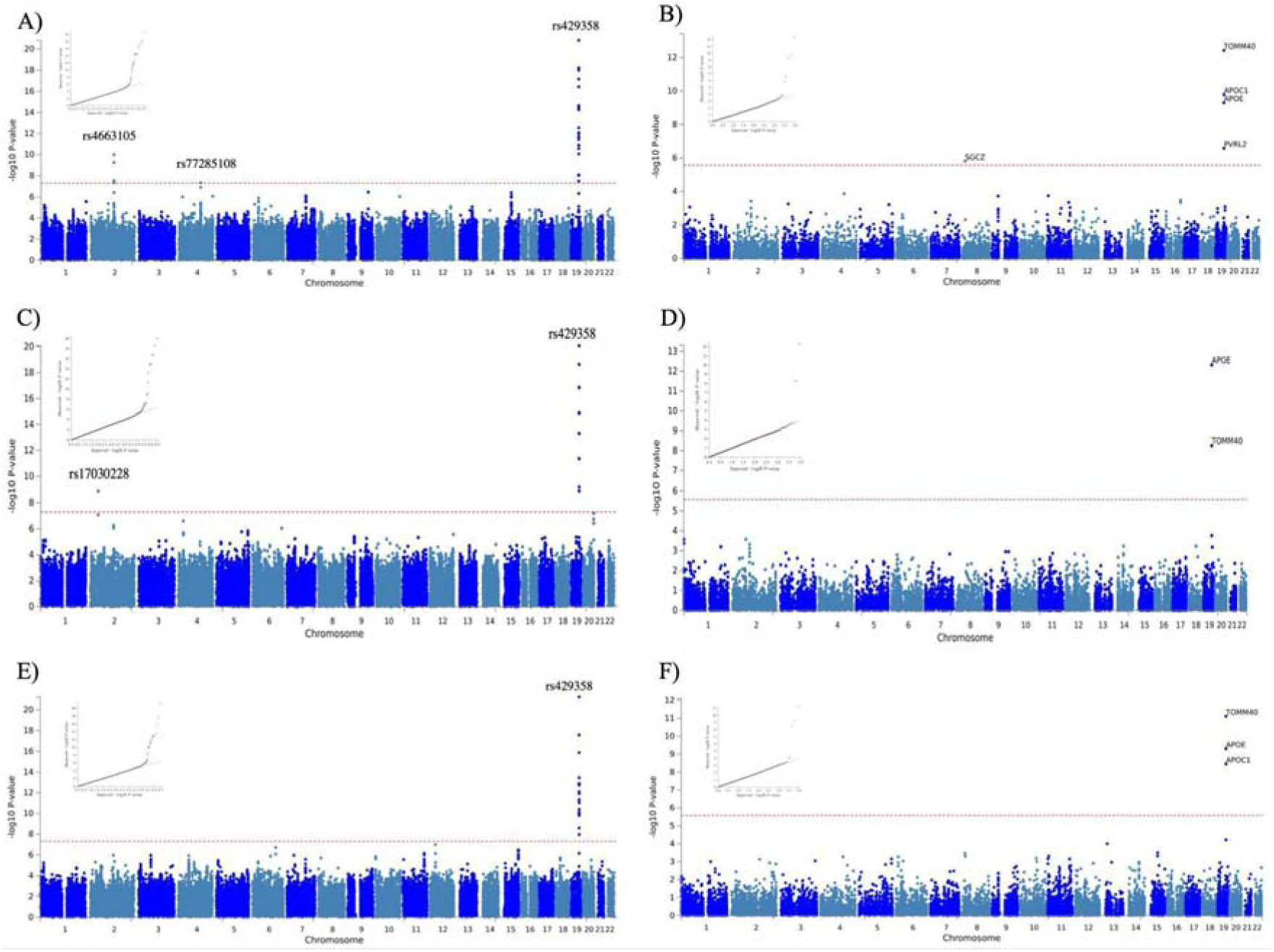
Manhattan and QQ plots of SNP-based and Gene-Based female-specific GWAS results of neuropathological features of AD (n□=2660). Dotted red lines represent the threshold for genomewide significance (P <5×10^−8^) and Bonferroni correction for the gene-based analyses. A) Neuropathologically-confirmed AD case-control female-specific GWAS. B) Gene-based analysis for neuropathologically-confirmed AD female-specific case-control sample. C) Braak stage female-specific GWAS. D) Gene-based analysis for Braak stage in females. E) NP score female-specific GWAS. F) Gene-based analysis for NP score in females.

**Table 1:**
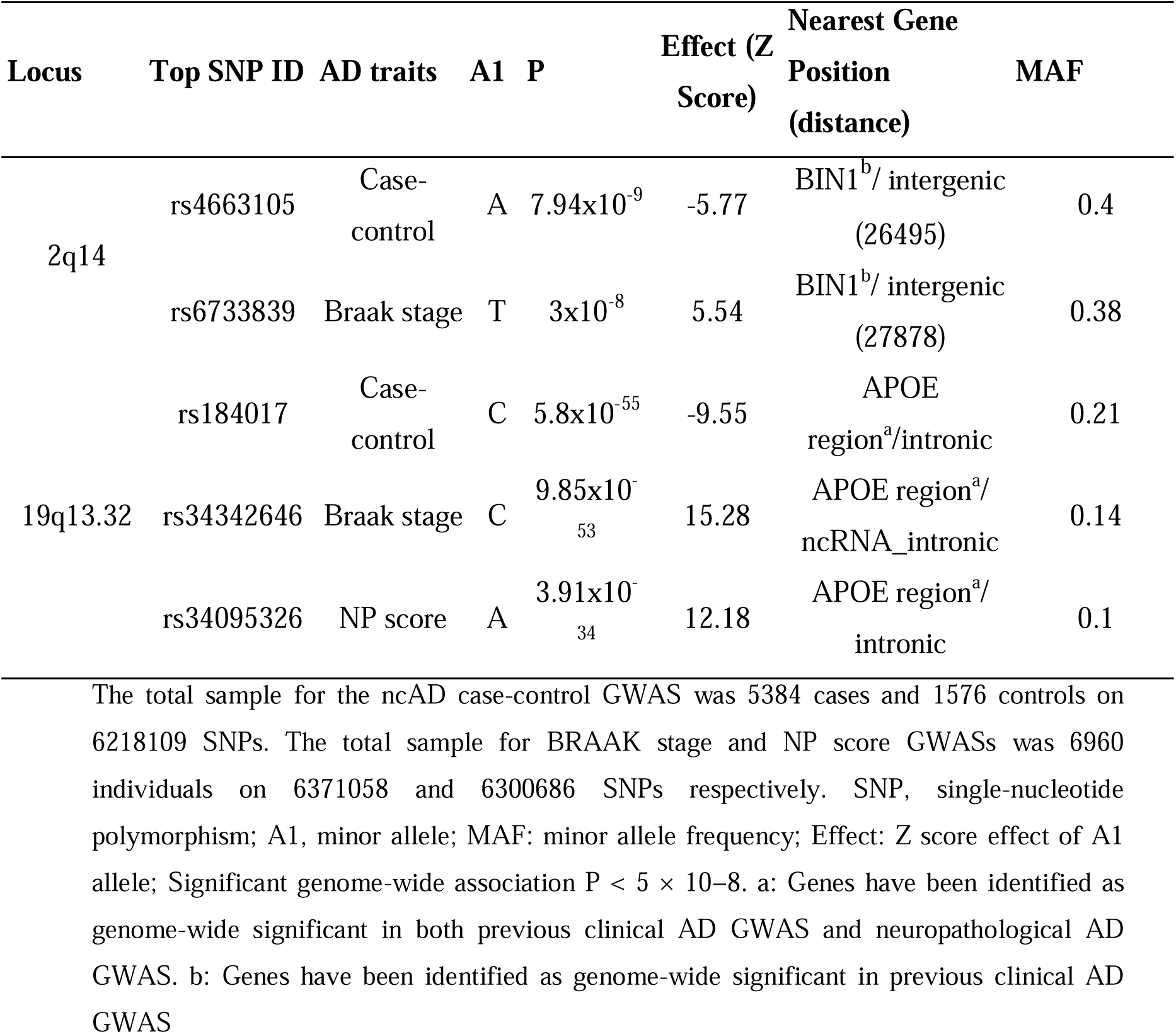
Genome-wide association study of neuropathological AD traits.

In the gene-based analyses, TOMM40, PVRL2, and APOC1 genes were significantly associated with all three studied neuropathology-based AD-related phenotypes (Figures 2B, 2D, 2F). The APOE gene was also significant in the Braak stage meta-analysis (Figure 1D), while two novel genes, OPCML and CDH4, outside of the 19q13.32 region were significant in the NP score GWAS (Figure 1E).

### Sex-specific genome-wide association studies of AD neuropathology

Next, aiming to investigate sex-specific genetic associations with AD neuropathology traits, we performed sex-stratified GWAS after merging seven datasets with available sex information (Supplementary Table S2) This resulted in a total of 2,366 males and 2,660 females. As expected, for both sexes, the top locus in all three AD neuropathology phenotypes that we studied (ncAD, Braak stage, NP score) was 19q13.32 with the top SNPs located in the APOE region (Table 2, Figure 2A, 2C, 2E, Supplementary Figure 1A, 1C, 1E). No additional significant loci were identified in the male specific GWAS for any of the three phenotypes (Table 2). However, three additional loci exceeded the genomewide significance level in the female-specific GWAS for Braak stage and ncAD: 2q14 close to the BIN1 gene and 4q27 close to QRFPR were significantly associated with ncAD in females (top SNPs: (2q14) rs4663105, p-value=1.01×10^−10^, Z=-6.47; (4q27) rs77285108, p-value=4.67×10^−8^, Z=5.46) (Figure 2C, 2E, Supplementary Figure 4A-B). Furthermore, rs17030228 close to the LOC102723854 was significant in the female-specific Braak stage GWAS (top SNP:rs17030228, p-value=8.5×10^−8^, Z=-6.08) (Table 2, Figure 2C, Supplementary Figure 4C). In gene-based analysis, genes in the 19q13.32 region (APOE region) were significant for all three AD neuropathology traits for both sexes, while the SGCZ gene was significant in female specific ncAD (Figure 2B, 2D, 2F, Supplementary Figure 1B, 1D,1F).

**Table 2:**
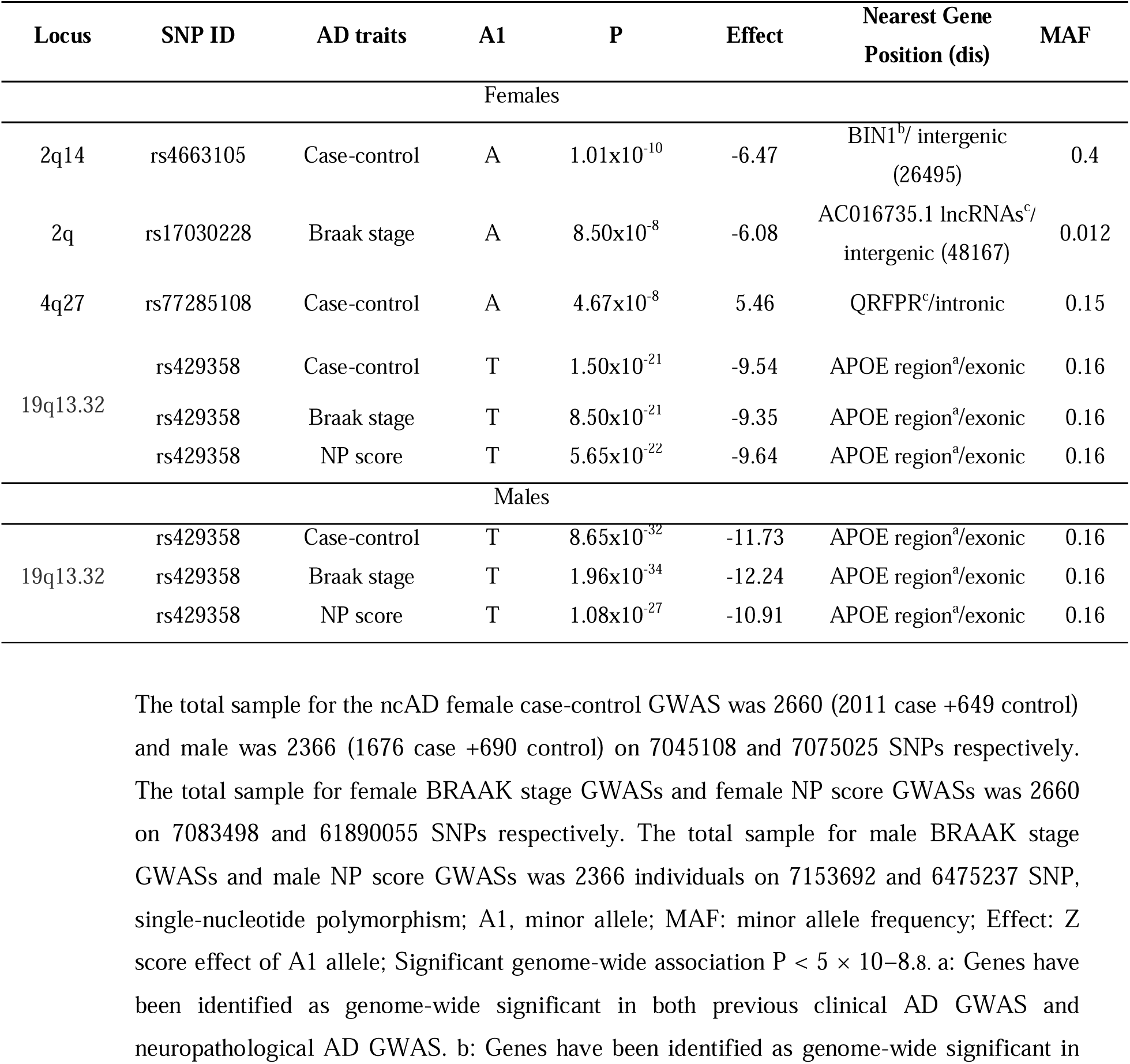
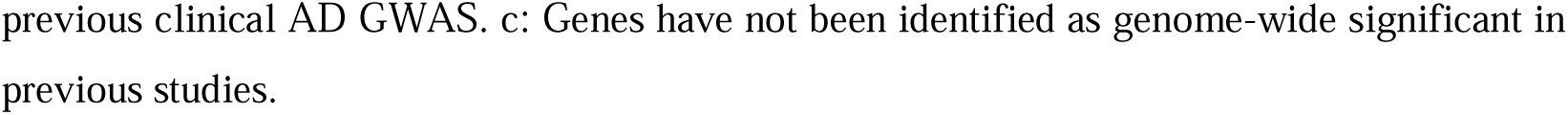
Sex-specific genome-wide association study of neuropathological traits in AD.

| Locus | SNP ID | AD traits | A1 | P | Effect | Nearest Gene Position (dis) | MAF |
| --- | --- | --- | --- | --- | --- | --- | --- |
| Females |  |  |  |  |  |  |  |
| 2q14 | rs4663105 | Case-control | A | $1.01 \times 10^{-10}$ | -6.47 | BIN1<sup>b</sup>/ intergenic (26495) | 0.4 |
| 2q | rs17030228 | Braak stage | A | $8.50 \times 10^{-8}$ | -6.08 | AC016735.1 lncRNAs<sup>c</sup>/ intergenic (48167) | 0.012 |
| 4q27 | rs77285108 | Case-control | A | $4.67 \times 10^{-8}$ | 5.46 | QRFPR<sup>c</sup>/intronic | 0.15 |
| 19q13.32 | rs429358 | Case-control | T | $1.50 \times 10^{-21}$ | -9.54 | APOE region<sup>a</sup>/exonic | 0.16 |
| | rs429358 | Braak stage | T | $8.50 \times 10^{-21}$ | -9.35 | APOE region<sup>a</sup>/exonic | 0.16 |
| | rs429358 | NP score | T | $5.65 \times 10^{-22}$ | -9.64 | APOE region<sup>a</sup>/exonic | 0.16 |
| Males |  |  |  |  |  |  |  |
| 19q13.32 | rs429358 | Case-control | T | $8.65 \times 10^{-32}$ | -11.73 | APOE region<sup>a</sup>/exonic | 0.16 |
| | rs429358 | Braak stage | T | $1.96 \times 10^{-34}$ | -12.24 | APOE region<sup>a</sup>/exonic | 0.16 |
| | rs429358 | NP score | T | $1.08 \times 10^{-27}$ | -10.91 | APOE region<sup>a</sup>/exonic | 0.16 |

The total sample for the ncAD female case-control GWAS was 2660 (2011 case +649 control) and male was 2366 (1676 case +690 control) on 7045108 and 7075025 SNPs respectively. The total sample for female BRAAK stage GWASs and female NP score GWASs was 2660 on 7083498 and 61890055 SNPs respectively. The total sample for male BRAAK stage GWASs and male NP score GWASs was 2366 individuals on 7153692 and 6475237 SNP, single-nucleotide polymorphism; A1, minor allele; MAF: minor allele frequency; Effect: Z score effect of A1 allele; Significant genome-wide association $P < 5 \times 10^{-8}$ . a: Genes have been identified as genome-wide significant in both previous clinical AD GWAS and neuropathological AD GWAS. b: Genes have been identified as genome-wide significant in
previous clinical AD GWAS. c: Genes have not been identified as genome-wide significant in previous studies.

### Post-GWAS analysis for AD neuropathology

To identify tissue specificity of our GWAS findings related to AD neuropathology, we performed tissue enrichment analysis by MAGMA. Notably, a significant association was found between NP score for females and genes expressed in the ovary tissue. (Figure 3). In the gene-set analysis, we observed one significantly enriched gene set for the case-control ncAD: “formation of anatomical boundary” (P_bon_=0.04). In the gene-set enrichment analysis of the gene-based results of the male case-control ncAD, we identified one significantly enriched GO term “histone pre mRNA 3 end processing complex” (P_bon_=0.04). We also observed one significant gene-set using the NP score in males, specifically “positive regulation of mitochondrial calcium ion concentration” (P_bon_=0.04). In the case of females, two gene-sets were significantly enriched when studied the NP score: “regulation of luteinizing hormone secretion” (P_bon_=0.00001) and “positive regulation of gonadotropin secretion” (P_bon_=0.0017). Furthermore, we found the gene-set “positive regulation of receptor catabolic process” (P_bon_=0.026) to be significantly enriched when studied genes identified in Braak stage at females, while in the female case-control ncAD analysis, we found one significant gene-set “fibroblast migration” (P_bon_=0.02) (Supplementary Table S3).

**Figure 3:**
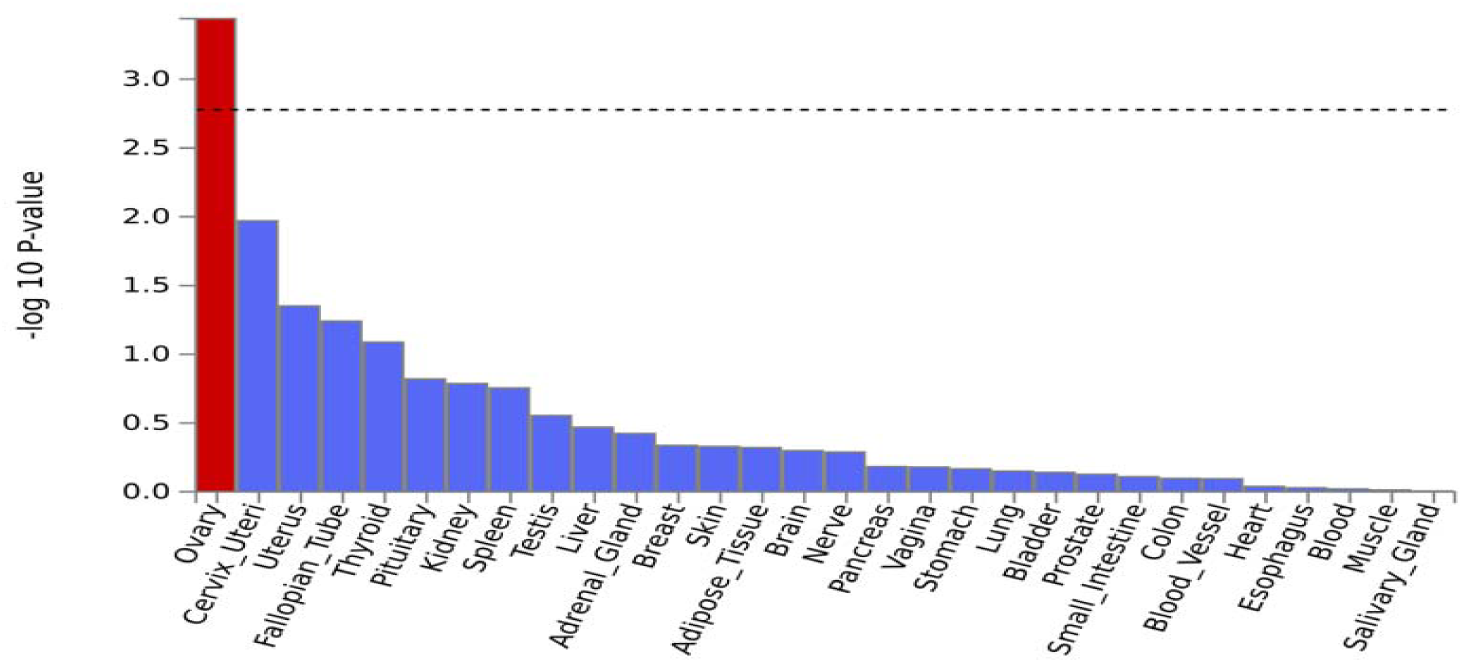
Post-GWAS analysis results for Tissue enrichment analysis. Tissue enrichment analysis for NP score GWAS results in females The analysis was performed in MAGMA using GTEx v8 RNA-seq data 30 general tissue types. With red are shown the significant results after multiple testing corrections.

To identify candidate genes whose genetically regulated expression is associated with neuropathological features of AD, we conducted a transcriptome-wide association study (TWAS – see Table 3 and Fig S6). This analysis identified nine protein-coding and two significant long non-coding RNA gene hits whose transcript expression was significantly associated with neuropathological features of AD. Notably, TWAS identified seven novel loci including genes (ST8SIA1 p-value=1.05×10-7; EIF2B2 p-value=1.00×10-6; NEUROD2 p-value=2.40×10-6; MRPL38 p-value=1.42×10-11; CSKMT p-value=3.50×10-8; CTXN2-AS1 p-value=9.21×10-12; LINC02458 p-value=2.50×10-7) and one novel gene in the APOE region (SYT5 p-value=8.10×10-9). These eight genes have not been implicated in previous AD-related GWAS or TWAS and are novel findings of this study.

**Table 3:**
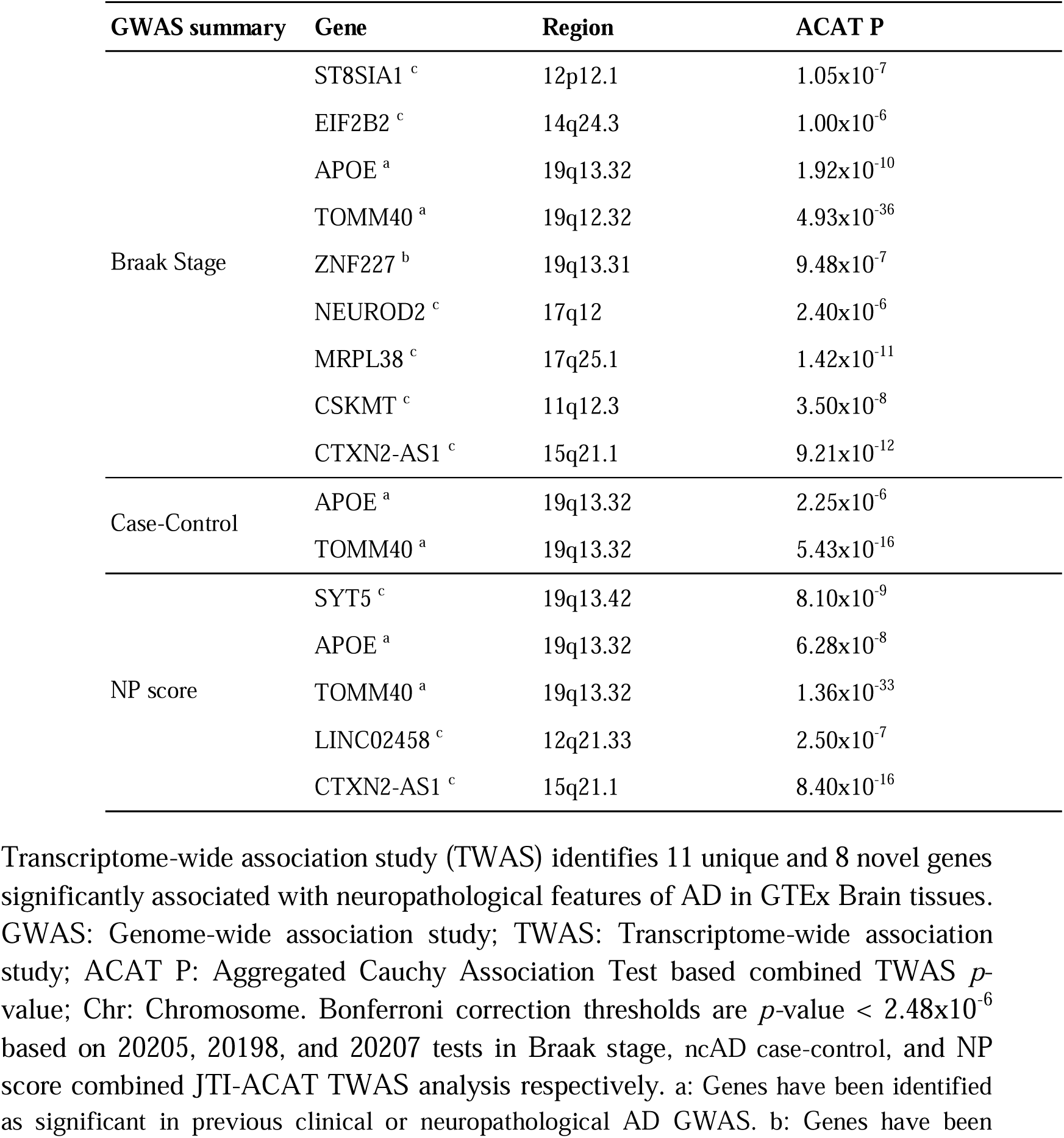
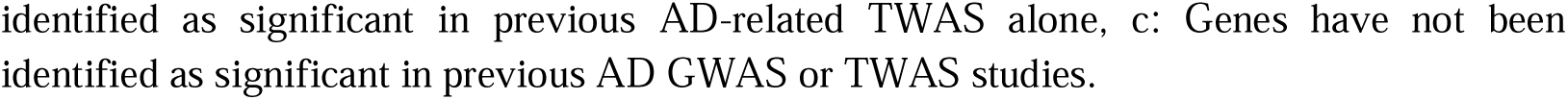
Transcriptome-wide association study of neuropathological traits in AD.

| GWAS summary | Gene | Region | ACAT P |
| --- | --- | --- | --- |
| Braak Stage | ST8SIA1 <sup>c</sup> | 12p12.1 | 1.05x10<sup>-7</sup> |
|  | EIF2B2 <sup>c</sup> | 14q24.3 | 1.00x10<sup>-6</sup> |
|  | APOE <sup>a</sup> | 19q13.32 | 1.92x10<sup>-10</sup> |
|  | TOMM40 <sup>a</sup> | 19q12.32 | 4.93x10<sup>-36</sup> |
|  | ZNF227 <sup>b</sup> | 19q13.31 | 9.48x10<sup>-7</sup> |
|  | NEUROD2 <sup>c</sup> | 17q12 | 2.40x10<sup>-6</sup> |
|  | MRPL38 <sup>c</sup> | 17q25.1 | 1.42x10<sup>-11</sup> |
|  | CSKMT <sup>c</sup> | 11q12.3 | 3.50x10<sup>-8</sup> |
|  | CTXN2-AS1 <sup>c</sup> | 15q21.1 | 9.21x10<sup>-12</sup> |
| Case-Control | APOE <sup>a</sup> | 19q13.32 | 2.25x10<sup>-6</sup> |
|  | TOMM40 <sup>a</sup> | 19q13.32 | 5.43x10<sup>-16</sup> |
| NP score | SYT5 <sup>c</sup> | 19q13.42 | 8.10x10<sup>-9</sup> |
|  | APOE <sup>a</sup> | 19q13.32 | 6.28x10<sup>-8</sup> |
|  | TOMM40 <sup>a</sup> | 19q13.32 | 1.36x10<sup>-33</sup> |
|  | LINC02458 <sup>c</sup> | 12q21.33 | 2.50x10<sup>-7</sup> |
|  | CTXN2-AS1 <sup>c</sup> | 15q21.1 | 8.40x10<sup>-16</sup> |

Transcriptome-wide association study (TWAS) identifies 11 unique and 8 novel genes significantly associated with neuropathological features of AD in GTEx Brain tissues. GWAS: Genome-wide association study; TWAS: Transcriptome-wide association study; ACAT P: Aggregated Cauchy Association Test based combined TWAS *p*-value; Chr: Chromosome. Bonferroni correction thresholds are *p*-value < 2.48x10<sup>-6</sup> based on 20205, 20198, and 20207 tests in Braak stage, ncAD case-control, and NP score combined JTI-ACAT TWAS analysis respectively. a: Genes have been identified as significant in previous clinical or neuropathological AD GWAS. b: Genes have been
identified as significant in previous AD-related TWAS alone, c: Genes have not been identified as significant in previous AD GWAS or TWAS studies.

**Table 4.**
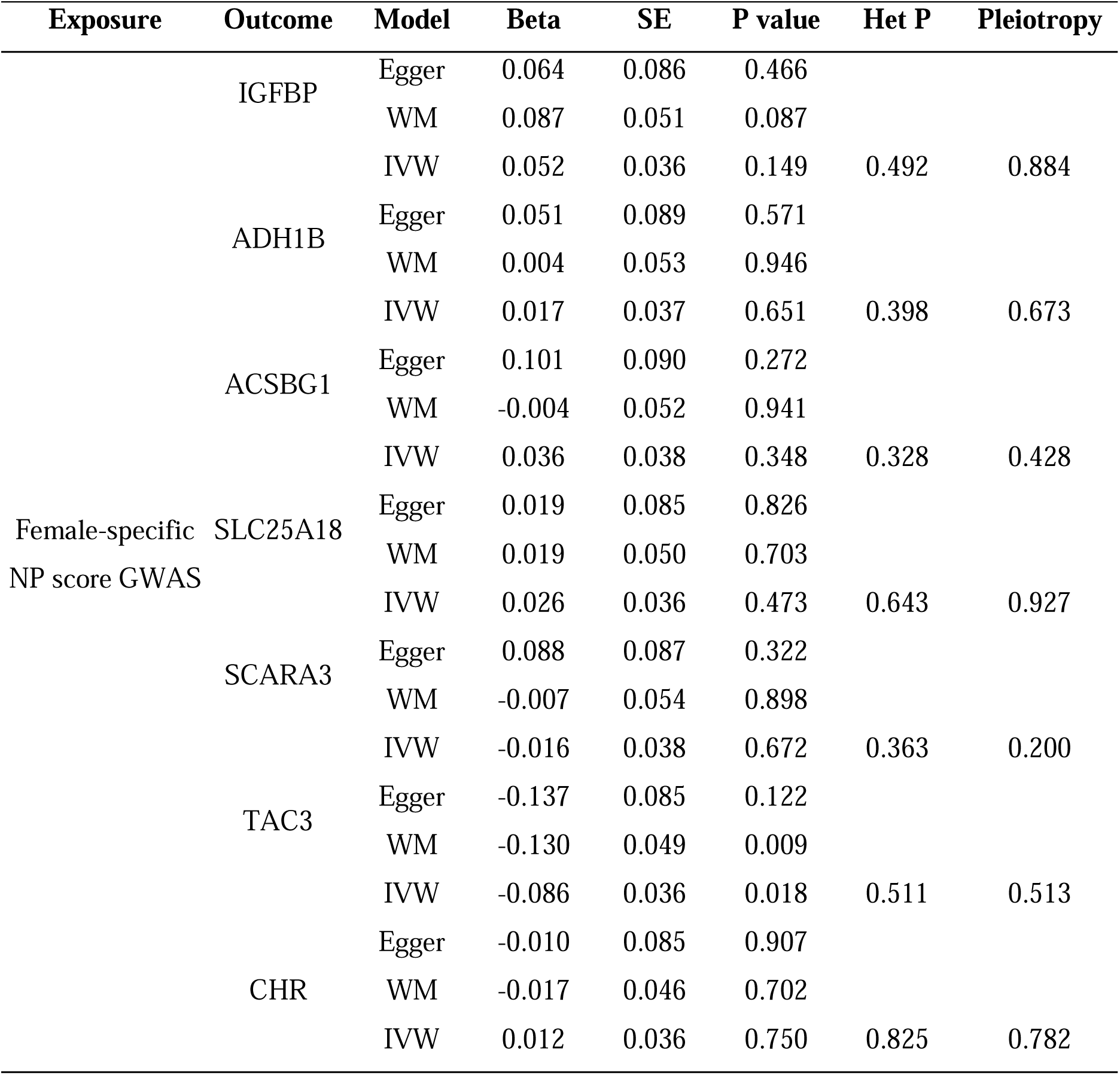
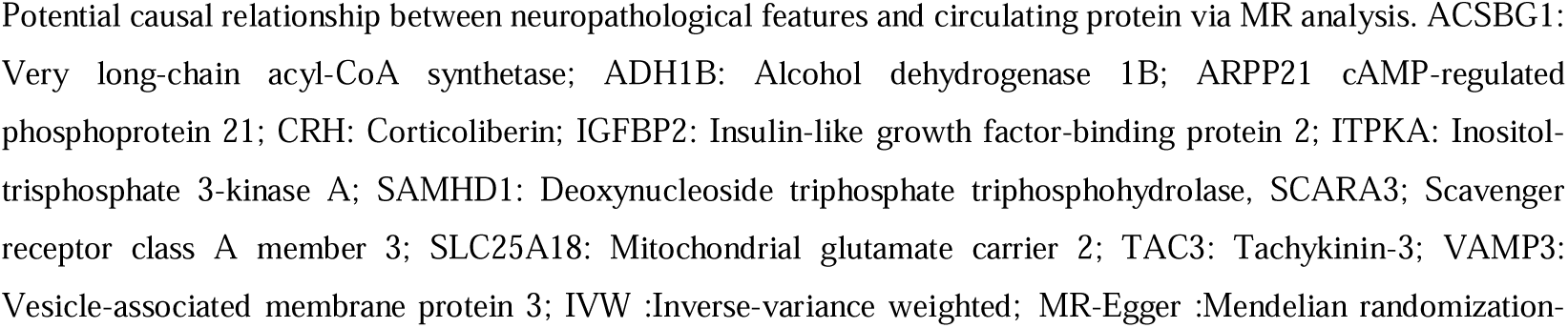
Mendelian randomization estimates causal relationship between Braak stage and circulating proteins.

### Identification of in-vivo biomarkers for AD neuropathology based on multi-omics and MR analysis

Finally, to explore biologically meaningful biomarkers associated with AD neuropathology traits, we employed an integrative analysis of multi-omics methods and MR. To that end, we explored i) differentially expressed genes (DEGs) using RNA seq data (Methods, Supplementary Table S4-5), ii) protein expression levels using the ROSMAP dataset, and iii) causal associations of blood plasma proteins from the INTERVAL study with AD neuropathology traits. To ensure the robustness of our results, we restricted all analyses to 683 biomarkers present in all three datasets. Figure 4 illustrates our workflow for in-vivo biomarker identification.

**Figure 4:**
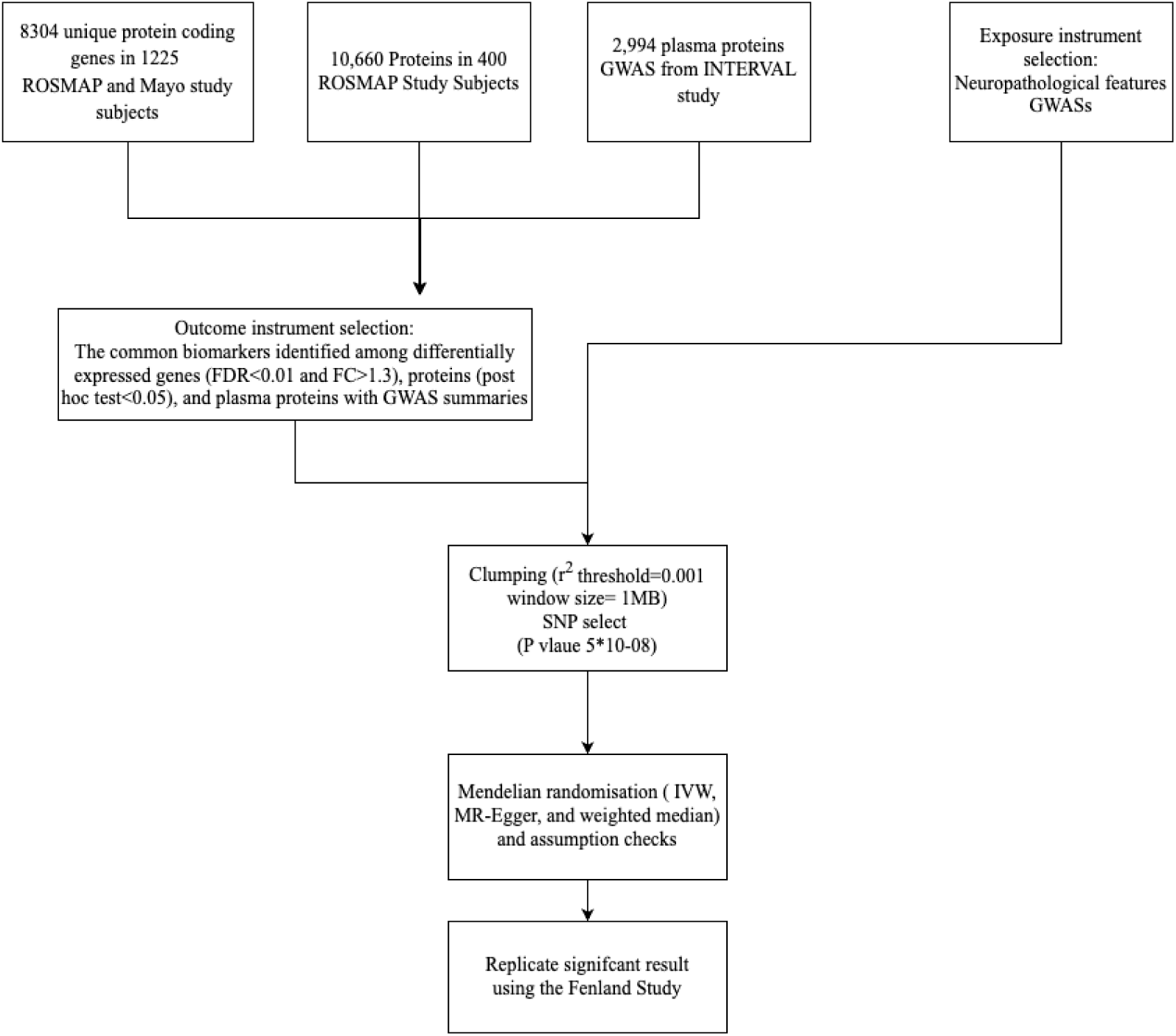
Overview of the pipeline for identification of in-vivo biomarkers using multi-omics and MR analysis. ROSMAP: Religious Orders Study and Memory and Aging Project; Mayo:Mayo Clinic Alzheimer’s Disease Research Center; IVW: Inverse-variance weighted; MR-Egger: Mendelian randomization-Egger and WM: weighted median; Interval study; The Fenland study;

In the differential gene expression analysis, the Mayo datasets, which included samples from the temporal lobe, we identified 142 genes with differential expression. Among these genes, 51 were downregulated, and 91 were upregulated (See Supplementary Table S4). Additionally, when considering ROSMAP cohorts with samples from the temporal lobe, we found 21 DEGs, including 12 downregulated and nine upregulated genes (Supplementary Table S5). Among these, 13 genes exhibited consistent regulation in at least two datasets, with seven showing downregulation and six showing upregulation, all in the same direction (Figure 5A).

**Figure 5:**
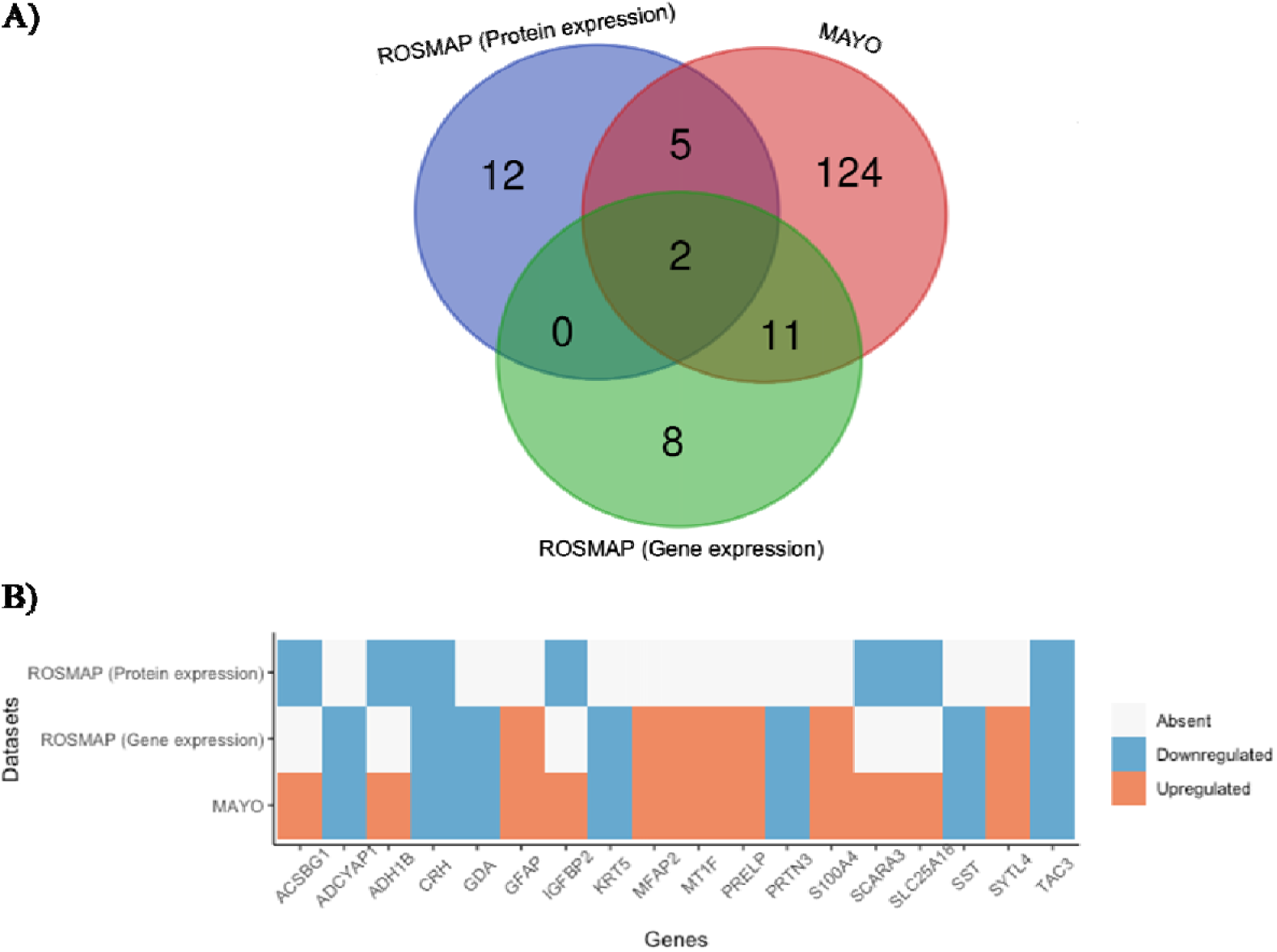
Identification of in-vivo biomarkers for AD neuropathology based on multi-omics analyses. A) Venn diagram illustrating significant differentially expressed genes and proteins and their overlap in the three datasets: MAYO, ROSMAP (gene expression) and ROSMAP (protein expression). B) Heatmap of the 18 genes that overlap in at least two datasets. With blue are shown the downregulated genes, red shows the upregulated genes, and white shows that the gene was either not significant or not present in the dataset.

Further analysis of protein expression revealed that 18 proteins exhibited a significant difference between the ncAD and control groups. Specifically, six proteins were significantly upregulated, while 12 proteins were significantly downregulated (Supplementary Table S6). Notably, seven of these proteins corresponded to genes that had previously shown significant changes in expression levels based on the aforementioned analysis of differentially expressed genes (Figure 5A-B).

We conducted MR (Mendelian Randomization) analysis on two separate samples to investigate the potentially causal relationship between neuropathological features and circulating protein. These circulating proteins overlap with those differentially expressed at both the protein and gene levels, as identified previously. We employed two-sample summary data MR methods, including IVW, MR-Egger, and weighted median. Our findings indicated a causal, positive relationship between Braak stage and ACSBG1 levels in blood (Beta: 0.065, P = 0.024 using IVW method), which was consistent with the alternative WM and Egger methods (Table 3). In sex-specific findings, the NP score in females showed a negative association with blood TAC3 levels (Beta: –0.085, P = 0.018, using the IVW method) which was also consistent with the alternative WM and Egger method. Seeking independent replication, we performed MR analysis using TAC3 GWAS data from the Fenland Study. We observed a similar directional trend in the relationship between NP score and TAC3 levels in females, although the association did not reach statistical significance. (Beta: –0.01, P = 0.44 using the IVW method). We could not explore ACSBG1 in the independent sample, because data were not available. Additional details can be found in Supplementary Table S7.

## Discussion

Integrating neuropathology, genomic, transcriptomic, and proteomic data we investigated the genetic basis of neuropathologically-confirmed AD (ncAD). We also pursued sex-specific analysis and sought to identify causal relationships and in-vivo biomarkers that could help track disease progression. Consistent with previous studies on clinically diagnosed AD, we find that the 19q and APOE regions yield the strongest signals of association also with ncAD. However, we also find novel loci that are associated with AD neuropathology and had not been previously found in clinical AD GWAS.

Our study of two AD-related neuropathology traits consistently showed SNPs located on chromosome 2q at the BIN1 locus to surpass genome-wide significance levels. This locus had not been identified by the previous neuropathology based GWAS^11^. However, BIN1 is one of the variants identified as associated with AD by the International Genomics of Alzheimer’s Project GWAS ^17^, and its significance should be emphasized since our work verified its implication in ncAD. BIN1 plays a prominent role in regulating endocytosis and synaptic vesicle trafficking, and it is implicated in the generation of amyloid beta and the propagation of Tau ^18–20^. This underscores its crucial role in the development of AD.

We also identified novel AD genes that surpassed genomewide significance levels in our association tests. The NP score GWAS identified significant genes on chromosome 11q (OPCML) and chromosome 20q (CDH4). OPCML (Opioid-Binding Protein/Cell Adhesion Molecule) belongs to the immunoglobulin protein superfamily and contributes to synaptogenesis in the brain ^21^. It has been implicated in AD based on previous GWAS studies in the Dutch population^22^. Additionally, GWAS analyses investigating the increased risk of AD in viral encephalitis have also revealed an association with OPCML^23^. CDH4 had not been previously implicated via AD GWAS. However, a study by Nazarian et al. (2021) found that CDH4 was almost significant with a P value of 3.44×10^−6^ in female-specific analysis^24^. CDH4 encodes the R-adherin protein, known for its involvement in cell migration and adhesion^25^. Previous animal studies suggest that this cadherin plays a crucial role in neuron development and the formation of vascular epithelial structures in the brain, potentially serving as a marker for endothelial injury in preclinical AD^26,27^. PHF21B, previously identified as a significant hit in neuropathology GWAS studies, was excluded from our analysis here due to high heterogeneity.

Our findings from the sex-specific GWAS identified significantly associated SNPs on chromosome 2q (BIN1 locus) and 4q (QRFPR) for case-control ncAD in females only providing new insights into the gender-specific genetic basis for AD progression. In a recent sex-specific clinical GWAS for AD, BIN1 was also identified as having a female-specific association^28^. Another investigation, estimating hazard ratios, revealed that BIN1 contributes to a higher risk in females compared to males^29^. Moreover, GTEx RNAseq analysis has previously underscored the sex-heterogeneous effect of BIN1 in brain tissue^30,31^. These findings suggest that the effects of BIN1 may be sex-dependent, particularly in the context of pathological AD. On the other hand, QRPFR (GPR103) acts as a receptor highly expressed in the brain for the orexigenic neuropeptide, influencing the regulation of feeding behavior and circadian rhythms^32,33^. Interestingly, disrupted circadian rhythms are linked to AD development, suggesting a potential role of QRPFR in AD pathology^34,35^. Additionally, QRPFR exhibits a neuroprotective effect, and its expression is reduced in AD due to amyloid-beta and tau pathology. The intra-hippocampal administration of orexin was shown to mitigate learning and memory impairment, highlighting its potential therapeutic role in AD^36^. Our Braak stage GWAS for females, implicates a novel long non-coding RNA (lncRNA), AC016735.1. Although studies that have explored this lncRNA in the context of AD are limited, emerging evidence indicates aberrant expression of lncRNAs in AD progression^37,38^. Our female-specific gene-based association study with case-control ncAD also identified SGCZ as a novel significant gene. SGCZ plays a role in forming the sarcoglycan complex and exhibits gender-biased expression levels in the brain, as observed in animal models^39^. Mutations in sarcoglycanopathy can lead to protein misfolding and aggregation, potentially contributing to the development of AD^40^. A single-cell analysis study revealed elevated expression of SGCZ in a subset of oligodendrocytes when comparing individuals with AD to those without the condition^41^. Finally, our identification of tissue-specific associations with AD in females, particularly within the ovarian region, presents a novel and intriguing insight, pointing to connections between sex hormones produced during the ovarian cycle and AD^42^. Intriguingly, hormonal therapy has been considered a potential treatment for reducing the risk of dementia^43^.

Through TWAS we identified eight novel genes as regulated by GWAS variants. Six genes (ST8SIA1, EIF2B2, NEUROD2, MRPL38, CSKMT and CTXN2-AS1) are associated with Braak stage and three (SYT5, LINC02458 and CTXN2-AS1) are associated with NP-score. ST8SIA1 gene encodes GD3 synthase involved in regulating amyloid-beta plaque load. EIF2B2 is implicated in brain protein synthesis, synaptic plasticity, and memory in a mouse model of AD ^44^. Additionally, two lncRNA genes LINC02458 and CTXN2-AS1, may play functional roles such as acting as a decoy or scaffold, influencing processes like amyloid beta aggregation, tau hyperphosphorylation, and the interaction of key enzymes in AD^45^. Further biological experimentation is warranted to provide additional support for the implication of genes in AD pathology.

The finding of a causal, negative relationship between NP score and TAC3 levels in the blood of females is particularly interesting although we were not able to replicate in the independent dataset that was available to us, and further replication should be sought. Nevertheless, TAC3 has been previously implicated in AD and is responsible for encoding neurokinin B (NKB), a neuropeptide that regulates various physiological processes, including calcium regulation and toxicity of Aβ process related to AD^46^. Additionally, TAC3, the tachykinin neuropeptide, has been shown to reduce the toxicity of Aβ, a hallmark protein associated with AD^47^. Our results suggest that the downregulation of TAC3 may be reflected in changes in protein levels in the blood, similar to what has been observed in CSF, and may serve as a potential biomarker or treatment target for the extent of neuropathological changes in AD^48^. Importantly, TAC3 is one of the genes that have been previously identified as a druggable target based on genomic information^49^. Concerning the potential utilization of ACSBG1 blood levels as a biomarker, ACSBG1 is prominently concentrated in the brain, primarily within astrocytes^50^. In vitro studies have demonstrated that ACSBG1 knockdown leads to reduced lipid oxidation, closely associated with AD ^51,52^ It is important to note that further research with larger sample sizes is required to confirm and expand upon our findings.

There are several limitations to the results discussed above that should be taken into consideration. First, the GWAS of protein biomarkers derived from blood samples, may not reflect the expression patterns in the brain, where AD pathology primarily occurs. Here, through MR analysis we did attempt to also explore causal relationships between AD neuropathological changes and the tested protein biomarkers, but the replicated results exhibited only marginal significance. Furthermore, while our findings hint at potential blood-based biomarkers for the neuropathological features of AD, the replicated results only exhibit a similar directional trend.

In summary, following a multifaceted and integrated multi-omics and neuropathology-based approach we revealed important insights into the genetic basis of true AD in both genders highlighting the potential significance of tissue-specific associations in understanding the etiology of the disease. The identification of novel SNPs and genes associated with AD neuropathology further contributes to our understanding of its molecular mechanisms. Further research is warranted to validate our findings in larger, independent cohorts, explore expression patterns in brain tissue and CSF samples and determine the clinical utility of our identified biomarkers for diagnosis, prognosis, and treatment of AD.

## Methods

### Datasets

Here, we put together a sample of 6,960 individuals integrating 13 large-scale genetic, clinical, and neuropathology datasets from multiple sources (Supplementary Materials, Supplementary Table S1). We included summary statistics datasets as described in Beecham et al^11^. along with additional individual-level datasets, to increase the sample size compared to the prior Alzheimer’s Disease Genetics Consortium (ADGC) study (full details provided in Supplementary Table S1). The expanded datasets included collaborative studies within ADGC, namely: 1) Alzheimer’s Disease Center (ADC) (with an increased sample size 1,074), and 2) Religious Orders Study and Memory and Aging Project (ROSMAP) (with an increased sample size 181). Furthermore, we included data from The Harvard Brain Tissue Resource Center (HBTRC) study (N=430) and the Alzheimer’s Disease Neuroimaging Initiative (N=71).

Samples were assessed for neuropathological changes using the guidelines recommended by the National Institute on Aging (NIA) and the Ronald and Nancy Reagan Institute of the Alzheimer’s Association. Criteria for ncAD cases and controls were based on CERAD scores for neuritic plaques (NPs) and Braak staging scores for tau neurofibrillary tangles (NFTs)^53,54^. For ncAD, a NP score of moderate/frequent and Braak stage III–VI were used, while for controls, a NP score of sparse and Braak stage less than II, or a NP score of none and Braak stage less than IV were used.

### Genomewide Association Studies

#### Genotyping and quality control process

The genotyping platforms that were used to assay samples in each cohort can be found in Supplementary Table S1. Quality control per dataset was performed as described ^55^. Briefly, samples with low call rate 98%, excessive heterozygosity rate 0.2, genomic sex discrepancy with reported sex, and formation of pairs with relatedness 0.4, were excluded from the downstream analyses. Variant-level quality control was performed to exclude markers with call rate 95%, and Hardy-Weinberg equilibrium p-value 10^−6^. To identify samples with European ancestry, Principal Component Analysis (PCA) was performed with EIGENSTRAT^56^ using 1000 Genomes as reference. Imputation on each dataset was performed via IMPUTE2 with 1000 Genomes as reference panel^57^ using data phased by SHAPEIT ^58^.

#### Genetic association, meta-analysis

In each dataset association tests were performed for three phenotypes; neuropathology-confirmed case/control (binary), Braak stage, and the NP score (ordinal) through PLINK ^59^ using the appropriate regression model and including the first three Principal Components (PCs) as covariates to adjust for population structure. Only variants with minor allele frequency > 0.01 and info score > 0.7 were included in the analyses. Following all quality controls in our study of AD neuropathological traits, a final meta-analysis was conducted using a total of 6,960 samples (Supplementary Table S1). Fixed-effects meta-analysis was performed using METAL^60^, and variants with heterogeneity (Cochran’s Q test P<0.05) and present in less than half of the subjects were removed.

#### Sex-specific GWAS

Seven of the analyzed datasets also had sex information available for part of their samples (Supplementary Table S2). We were thus able to perform sex specific GWAS. For these analyses quality control and imputation was followed as described above and for each dataset separately. The sex specific GWAS for the three phenotypes was performed through PLINK including the first three PCs and age as covariates. Sex-specific meta-analysis of 2,660 females and 2,366 males was performed as described earlier (Methods).

#### Post GWAS analyses

Gene-based analyses were conducted within FUMA^61^ using MAGMA^62^, and the significance level was calculated after Bonferroni correction accounting for the tested genes. For tissue expression analysis, we employed MAGMA, which utilized gene expression data per tissue from GTEx v8 RNA-seq data, with significance set at P<1.67 × 10^−3^ ^30^ (after Bonferroni correction for 30 tests). To determine whether the genetic findings associated with neuropathological features converged on functional gene sets and pathways, we performed gene set analyses using MAGMA. Gene sets included in the analyses were the “GO terms” from Msigdb v7.0 and included genes as detected by MAGMA gene-based test. P_bon_<0.05 was set as the significance threshold for gene-set analysis accounting for multiple-tests. Additionally, a transcriptome-wide association study (TWAS) was performed using the Joint-Tissue Imputation (JTI) method ^63^ with a goal of identifying genes regulated by disease-associated variants. Precomputed gene expression weights from 14 brain tissues in GTEx v8 were used to accomplish transcriptomic imputation and estimate a gene’s association with neuropathological features of AD. Gene associations were considered significant if the TWAS *p*-value was less than 0.05 divided by the number of genes tested per tissue. Additionally, we combined TWAS *p*-values from multiple tissues using the Aggregated Cauchy Association Test (ACAT) method ^64^ and performed the Bonferroni method to control for multiple tests.

### Identification of in-vivo biomarkers for AD

Next, we utilized integrative analysis of multi-omics methods and Mendelian Randomization (MR) to identify causal associations between AD neuropathological features and biologically meaningful biomarkers. To that end we integrated data from three different sources: 1) RNA-seq from ROSMAP and Mayo Clinic Alzheimer’s Disease Research Center (MAYO) 2) proteomics from ROSMAP 3) plasma protein GWAS summary statistics from INTERVAL study ^65^. Both the RNA-seq and proteomics datasets represent a subset of the samples included in the GWAS and are utilized to identify biomarkers with clinical significance.

#### Differential Gene Expression Analysis

For our differential gene expression analysis, datasets were acquired from ROSMAP and MAYO, which can be accessed at https://www.synapse.org. Data on a total of 583 subjects from six cohorts were extracted from this Portal. Mayo study, investigating the temporal cortex, had a sample size of 160 subjects comprising 82 AD and 78 controls. ROSMAP study investigated the Dorsolateral prefrontal cortex DLPFC, with a sample size of 423 subjects comprising 222 AD and 201 controls. Gene Expression (RNA seq) was preprocessed and analyzed as previously described^66^. Briefly, raw reads underwent trimming with Trimmomatic, followed by mapping to the human reference genome GRCh38 using STAR. Subsequently, gene expression quantification, normalization (trimmed mean of M-values), filtering (retaining genes with over 1 count per million in at least 30% of samples), and quality control (Picard) were performed. Then, we utilized the DESeq2 R package^67^ to detect Differentially Expressed Genes (DEGs) at the transcript level. The gene counts length and abundance obtained through tximport were analyzed using DESeq2. Significance cutoffs were applied to select genes with a P_FDR_□<□0.01 and a fold change (FC)□>□1.3.

#### Differential Protein Expression Analysis

The ROSMAP brain samples of 400 individuals, were obtained from the AD Knowledge Portal (https://www.synapse.org/#!Synapse:syn17015098) and subjected to TMT peptide labeling. The tissue homogenization and TMT mass spectrometry processing were conducted using the methods previously described ^68^. We included proteins in the analysis only if they met the criteria of a q-value < 0.01 and a protein missing rate < 50%. The identification of differentially expressed proteins involved performing one-way ANOVA, followed by Tukey’s Honest Significant Difference post hoc test, across control, mild cognitive impairment (MCI), and AD cases.

#### Mendelian Randomization (MR) Analysis

Two-sample MR analysis was conducted to examine the potential causal relationship between AD neuropathology traits and circulating protein abundances (generated by Sun et al. 2018; INTERVAL study). The INTERVAL study involved 3,301 individuals of European descent and conducted GWAS on the levels of 2,994 plasma proteins. Significant SNPs (p< 1□×□10^−5^) in the neuropathological AD GWAS summary statistics were considered for exposure variables. In an ideal scenario, MR analysis relies on genome-wide SNPs (P < 5×10^−8^). However, in previous MR studies with limited genome-wide SNPs, a relatively lenient threshold for genetic instruments has been employed ^69^. LD-independent SNPs were identified with the clump function in PLINK with r2 threshold of 0.001 and a window size of 1□Mb using 1000 Genomes European samples as reference. The next analyses were performed with the TwosampleMR R package ^70^. Inverse-variance weighted (IVW) regression with a multiplicative random effects model was conducted as the primary causal inference. While two additional methods, MR-Egger and weighted median (WM) were also applied to validate the results and test for pleiotropy. Additionally, Cochran’s Q test was applied to test for heterogeneity and sensitivity analysis was performed using leave-one-out analysis. Finally, the Fenland Study was used to replicate significant results ^71^.

## Data Availability

All data produced in the present work are contained in the manuscript

## References

1. Arvanitakis, Z., Shah, R. C. & Bennett, D. A. Diagnosis and Management of Dementia: Review. JAMA – J. Am. Med. Assoc. (2019) doi:10.1001/jama.2019.4782.

2. Schneider, J. A., Arvanitakis, Z., Bang, W. & Bennett, D. A. Mixed brain pathologies account for most dementia cases in community-dwelling older persons. Neurology (2007) doi:10.1212/01.wnl.0000271090.28148.24.

3. Jellinger, K. A. & Attems, J. Challenges of multimorbidity of the aging brain: a critical update. Journal of Neural Transmission vol. 122 505–521 (2015).

4. Fischer, C. E. et al. Determining the impact of psychosis on rates of false-positive and false-negative diagnosis in Alzheimer’s disease. in Alzheimer’s and Dementia: Translational Research and Clinical Interventions (2017). doi:10.1016/j.trci.2017.06.001.

5. Medway, C. & Morgan, K. Review: The genetics of Alzheimer’s disease; putting flesh on the bones. Neuropathol. Appl. Neurobiol. 40, 97–105 (2014).

6. Reitz, C. Genetic loci associated with Alzheimer’s disease. Future Neurol. (2014) doi:10.2217/fnl.14.1.

7. Sonnen, J. A. et al. Ecology of the Aging Human Brain. Arch. Neurol. 68, 1049–1056 (2011).

8. Bennett, D. A., Wilson, R. S., Boyle, P. A., Buchman, A. S. & Schneider, J. A. Relation of neuropathology to cognition in persons without cognitive impairment. Ann. Neurol. 72, 599–609 (2012).

9. Bennett, D. A., Schneider, J. A., Wilson, R. S., Bienias, J. L. & Arnold, S. E. Neurofibrillary Tangles Mediate the Association of Amyloid Load with Clinical Alzheimer Disease and Level of Cognitive Function. Arch. Neurol. (2004) doi:10.1001/archneur.61.3.378.

10. Farfel, J. M. et al. Relation of genomic variants for Alzheimer disease dementia to common neuropathologies. Neurology (2016) doi:10.1212/WNL.0000000000002909.

11. Beecham, G. W. et al. Genome-Wide Association Meta-analysis of Neuropathologic Features of Alzheimer{\textquotesingle}s Disease and Related Dementias. {PLoS} Genet. 10, e1004606 (2014).

12. 2023 Alzheimer’s disease facts and figures. Alzheimer’s Dement. (2023) doi:10.1002/alz.13016.

13. Dumitrescu, L. et al. Sex differences in the genetic predictors of Alzheimer’s pathology. Brain (2019) doi:10.1093/brain/awz206.

14. Tan, C. H. et al. Polygenic hazard score: an enrichment marker for Alzheimer’s associated amyloid and tau deposition. Acta Neuropathol. (2018) doi:10.1007/s00401-017-1789-4.

15. Jansen, W. J. et al. Prevalence of cerebral amyloid pathology in persons without dementia: A meta-analysis. JAMA – J. Am. Med. Assoc. 313, 1924– 1938 (2015).

16. Sperling, R. A. et al. Toward defining the preclinical stages of Alzheimer’s disease: Recommendations from the National Institute on Aging-Alzheimer’s Association workgroups on diagnostic guidelines for Alzheimer’s disease. Alzheimer’s Dement. (2011) doi:10.1016/j.jalz.2011.03.003.

17. Lambert, J.-C. et al. Meta-analysis of 74,046 individuals identifies 11 new susceptibility loci for Alzheimer’s disease. Nat. Genet. 45, 1452–1458 (2013).

18. Miyagawa, T. et al. BIN1 regulates BACE1 intracellular trafficking and amyloid-β production. Hum. Mol. Genet. (2016) doi:10.1093/hmg/ddw146.

19. Crotti, A. et al. BIN1 favors the spreading of Tau via extracellular vesicles. Sci. Rep. (2019) doi:10.1038/s41598-019-45676-0.

20. De Rossi, P. et al. Neuronal BIN1 Regulates Presynaptic Neurotransmitter Release and Memory Consolidation. Cell Rep. (2020) doi:10.1016/j.celrep.2020.02.026.

21. Hashimoto, T., Maekawa, S. & Miyata, S. IgLON cell adhesion molecules regulate synaptogenesis in hippocampal neurons. Cell Biochem. Funct. (2009) doi:10.1002/cbf.1600.

22. Liu, F. et al. A genomewide screen for late-onset Alzheimer disease in a genetically isolated Dutch population. Am. J. Hum. Genet. (2007) doi:10.1086/518720.

23. Lehrer, S. & Rheinstein, P. H. RORB, an Alzheimer’s disease susceptibility gene, is associated with viral encephalitis, an Alzheimer’s disease risk factor. Clin. Neurol. Neurosurg. 233, 1–15 (2023).

24. Nazarian, A., Yashin, A. I. & Kulminski, A. M. Genome-wide analysis of genetic predisposition to Alzheimer’s disease and related sex disparities. Alzheimer’s Res. Ther. (2019) doi:10.1186/s13195-018-0458-8.

25. Kools, P., Vanhalst, K. & Van Roy, F. Assignment of cadherin-4 (R-cadherin, CDH4) to human chromosome band 20q13.3. Cytogenet. Cell Genet. (1999).

26. Redies, C., Engelhart, K. & Takeichi, M. Differential expression of N□ and R□cadherin in functional neuronal systems and other structures of the developing chicken brain. J. Comp. Neurol. (1993) doi:10.1002/cne.903330307.

27. Tarawneh, R. et al. Vascular endothelial-cadherin as a marker of endothelial injury in preclinical Alzheimer disease. Ann. Clin. Transl. Neurol. (2022) doi:10.1002/acn3.51685.

28. Eissman, J. M. et al. Sex-specific genetic architecture of late-life memory performance. Alzheimer’s Dement. 1–18 (2023) doi:10.1002/alz.13507.

29. Fan, C. C. et al. Sex-dependent autosomal effects on clinical progression of Alzheimer’s disease. Brain (2020) doi:10.1093/brain/awaa164.

30. Aguet, F. et al. The GTEx Consortium atlas of genetic regulatory effects across human tissues. Science (80-.). (2020) doi:10.1126/SCIENCE.AAZ1776.

31. Guen, Y. Le et al. Sex-heterogenous effect on Alzheimer’s disease risk at the BIN1 locus. Alzheimers. Dement. (2021) doi:10.1002/alz.053616.

32. Takayasu, S. et al. A neuropeptide ligand of the G protein-coupled receptor GPR103 regulates feeding, behavioral arousal, and blood pressure in mice. Proc. Natl. Acad. Sci. U. S. A. (2006) doi:10.1073/pnas.0602371103.

33. Ma, Z., Jiang, W. & Zhang, E. E. Orexin signaling regulates both the hippocampal clock and the circadian oscillation of Alzheimer’s disease-risk genes. Sci. Rep. (2016) doi:10.1038/srep36035.

34. Musiek, E. S., Xiong, D. D. & Holtzman, D. M. Sleep, circadian rhythms, and the pathogenesis of Alzheimer Disease. Exp. Mol. Med. (2015) doi:10.1038/EMM.2014.121.

35. Milán-Tomás, Á. & Shapiro, C. M. Circadian Rhythms Disturbances in Alzheimer Disease. Alzheimer Disease and Associated Disorders (2018) doi:10.1097/WAD.0000000000000243.

36. Chen, X. Y., Du, Y. F. & Chen, L. Neuropeptides exert neuroprotective effects in alzheimer’s disease. Frontiers in Molecular Neuroscience (2019) doi:10.3389/fnmol.2018.00493.

37. Li, D. et al. Insights into lncRNAs in Alzheimer’s disease mechanisms. RNA Biology (2020) doi:10.1080/15476286.2020.1788848.

38. Luo, Q. & Chen, Y. Long noncoding RNAs and Alzheimer’s disease. Clinical Interventions in Aging (2016) doi:10.2147/CIA.S107037.

39. Hou, H. et al. Postnatal developmental trajectory of sex-biased gene expression in the mouse pituitary gland. Biol. Sex Differ. (2022) doi:10.1186/s13293-022-00467-7.

40. Sandonà, D. & Betto, R. Sarcoglycanopathies: Molecular pathogenesis and therapeutic prospects. Expert Reviews in Molecular Medicine (2009) doi:10.1017/S1462399409001203.

41. Sadick, J. S. et al. Astrocytes and oligodendrocytes undergo subtype-specific transcriptional changes in Alzheimer’s disease. Neuron 110, 1788–1805.e10 (2022).

42. Jett, S. et al. Ovarian steroid hormones: A long overlooked but critical contributor to brain aging and Alzheimer’s disease. Frontiers in Aging Neuroscience (2022) doi:10.3389/fnagi.2022.948219.

43. Nerattini, M. et al. Systematic review and meta-analysis of the effects of menopause hormone therapy on risk of Alzheimer’s disease and dementia. Front. Aging Neurosci. 15, (2023).

44. Oliveira, M. M. et al. Correction of eIF2-dependent defects in brain protein synthesis, synaptic plasticity, and memory in mouse models of Alzheimer’s disease. Sci. Signal. (2021) doi:10.1126/scisignal.abc5429.

45. Asadi, M. R. et al. The Perspective of Dysregulated LncRNAs in Alzheimer’s Disease: A Systematic Scoping Review. Frontiers in Aging Neuroscience (2021) doi:10.3389/fnagi.2021.709568.

46. Russino, D., McDonald, E., Hejazi, L., Hanson, G. R. & Jones, C. E. The tachykinin peptide neurokinin b binds copper forming an unusual □CuII(NKB)2] complex and inhibits copper uptake into 1321N1 astrocytoma cells. ACS Chem. Neurosci. (2013) doi:10.1021/cn4000988.

47. Yankner, B. A., Duffy, L. K. & Kirschner, D. A. Neurotrophic and neurotoxic effects of amyloid β protein: Reversal by tachykinin neuropeptides. Science (80-.). (1990) doi:10.1126/science.2218531.

48. Pedrero-Prieto, C. M. et al. A comprehensive systematic review of CSF proteins and peptides that define Alzheimer’s disease. Clinical Proteomics (2020) doi:10.1186/s12014-020-09276-9.

49. Finan, C. et al. The druggable genome and support for target identification and validation in drug development. Sci. Transl. Med. (2017) doi:10.1126/scitranslmed.aag1166.

50. Fernandez, R. F. & Ellis, J. M. Acyl-CoA synthetases as regulators of brain phospholipid acyl-chain diversity. Prostaglandins Leukotrienes and Essential Fatty Acids (2020) doi:10.1016/j.plefa.2020.102175.

51. Qi, G. et al. ApoE4 Impairs Neuron-Astrocyte Coupling of Fatty Acid Metabolism. Cell Rep. (2021) doi:10.1016/j.celrep.2020.108572.

52. Astrocyte, Lipid Metabolism in Alzheimer’s Disease and Glioblastoma. J. Clin. Res. Med. (2022) doi:10.31038/jcrm.2022555.

53. Fillenbaum, G. G. et al. Consortium to Establish a Registry for Alzheimer’s Disease (CERAD): The first twenty years. Alzheimer’s and Dementia (2008) doi:10.1016/j.jalz.2007.08.005.

54. Braak, H. & Braak, E. Neuropathological stageing of Alzheimer-related changes. Acta Neuropathologica (1991) doi:10.1007/BF00308809.

55. Naj, A. C. et al. Genome-wide meta-analysis of late-onset Alzheimer’s disease using rare variant imputation in 65,602 subjects identifies novel rare variant locus NCK2[: The International Genomics of Alzheimer’s Project (IGAP). medRxiv (2021).

56. Price, A. L. et al. Principal components analysis corrects for stratification in genome-wide association studies. Nat. Genet. 38, 904–909 (2006).

57. Howie, B. N., Donnelly, P. & Marchini, J. A flexible and accurate genotype imputation method for the next generation of genome-wide association studies. PLoS Genet. (2009) doi:10.1371/journal.pgen.1000529.

58. Delaneau, O., Coulonges, C. & Zagury, J. F. Shape-IT: New rapid and accurate algorithm for haplotype inference. BMC Bioinformatics (2008) doi:10.1186/1471-2105-9-540.

59. Purcell, S. et al. PLINK: a tool set for whole-genome association and population-based linkage analyses. Am. J. Hum. Genet. 81, 559–75 (2007).

60. Willer, C. J., Li, Y. & Abecasis, G. R. METAL: Fast and efficient meta-analysis of genomewide association scans. Bioinformatics (2010) doi:10.1093/bioinformatics/btq340.

61. Watanabe, K., Taskesen, E., Van Bochoven, A. & Posthuma, D. Functional mapping and annotation of genetic associations with FUMA. Nat. Commun. (2017) doi:10.1038/s41467-017-01261-5.

62. de Leeuw, C. A., Mooij, J. M., Heskes, T. & Posthuma, D. MAGMA: Generalized Gene-Set Analysis of GWAS Data. PLoS Comput. Biol. (2015) doi:10.1371/journal.pcbi.1004219.

63. Zhou, D. et al. A unified framework for joint-tissue transcriptome-wide association and Mendelian randomization analysis. Nat. Genet. (2020) doi:10.1038/s41588-020-0706-2.

64. Liu, Y. et al. ACAT: A Fast and Powerful p Value Combination Method for Rare-Variant Analysis in Sequencing Studies. Am. J. Hum. Genet. (2019) doi:10.1016/j.ajhg.2019.01.002.

65. Sun, B. B. et al. Genomic atlas of the human plasma proteome. Nature (2018) doi:10.1038/s41586-018-0175-2.

66. Marques-Coelho, D. et al. Differential transcript usage unravels gene expression alterations in Alzheimer’s disease human brains. *npj Aging Mech*. Dis. (2021) doi:10.1038/s41514-020-00052-5.

67. Love, M. I., Huber, W. & Anders, S. Moderated estimation of fold change and dispersion for RNA-seq data with DESeq2. Genome Biol. (2014) doi:10.1186/s13059-014-0550-8.

68. Ping, L. et al. Global quantitative analysis of the human brain proteome and phosphoproteome in Alzheimer’s disease. Sci. Data (2020) doi:10.1038/s41597-020-00650-8.

69. Choi, K. W. et al. Assessment of bidirectional relationships between physical activity and depression among adults a 2-sample Mendelian randomization study. JAMA Psychiatry (2019) doi:10.1001/jamapsychiatry.2018.4175.

70. Hemani, G. et al. The MR-base platform supports systematic causal inference across the human phenome. Elife (2018) doi:10.7554/eLife.34408.

71. Pietzner, M. et al. Mapping the proteo-genomic convergence of human diseases. Science (80-.). (2021) doi:10.1126/science.abj1541.

